# Time of day influences psychophysical measures in women with burning mouth syndrome

**DOI:** 10.1101/2021.02.03.21251110

**Authors:** Janell S. Payano Sosa, Joyce T. Da Silva, Shana A.B Burrowes, Soo Y. Yoo, Michael L. Keaser, Timothy F. Meiller, David A. Seminowicz

## Abstract

Burning mouth syndrome (BMS) is a chronic orofacial pain condition that mainly affects postmenopausal women. BMS type I patients report little to no spontaneous pain in the morning and increases in pain through the day, peaking in the afternoon. Quantitative sensory testing (QST) findings from BMS type 1 patients are inconsistent as they fail to capture this temporal variation. We examined how QST in BMS type 1 compared to healthy participants was affected by time of day. QST of the face and forearm included warmth detection threshold (WDT), cold detection threshold (CDT), and heat pain thresholds (HPT), ratings of suprathreshold heat, and pressure pain thresholds (PPT), and was performed twice: once in the morning and once in the afternoon. BMS patients reported: no differences in morning and afternoon comparisons of the orofacial region between groups and no differences in PPTs in either orofacial or arm regions between groups; lower within group PPTs of the masseter in the afternoon; higher WDTs, and lower CDTs of the forearm in the morning compared to healthy participants; lower CDTs and higher pain intensity ratings to heat stimuli at low temperatures on the forearm in the afternoon compared to healthy participants. The findings indicate that compared to healthy participants, BMS Type I patients had altered thermal sensitivity, which depended on body area tested (forearm vs orofacial region), and higher pain sensitivity, which was slightly more pronounced in the afternoon plausibly due to a hypervigilance.

**Perspective:** Somatosensory profiles in BMS type 1 can be affected by time of day, possibly as an effect of spontaneous burning pain that increases throughout the day. Our findings highlight the cyclical effect of BMS pathophysiology across a day, which may further help clinicians choose appropriate treatment strategies for BMS patients.

## Introduction

Burning Mouth Syndrome (BMS) is a chronic orofacial pain condition that mainly affects post-menopausal women^2, 3, 5, 6, 13, 19^. The most prevalent symptom of BMS is burning pain in the oral mucosa including the palate, inside lip, and the tip and anterior two-thirds of the tongue^1, 11^. However, the affected area of the oral mucosa is clinically normal^1, 11^. Therefore, in the absence of clear pathology in the oral mucosa, central mechanisms have been suggested to, at least in part, explain the spontaneous burning pain of BMS and the presence of pain in other body regions ^4, 9, 12^.

Somatosensory functions in people affected by BMS can be determined psychophysically using quantitative sensory testing (QST) ^14^. Previous QST studies have reported mixed results with some reporting BMS patients have increased sensitivity of the orofacial region to thermal stimuli relative to healthy participants (supplemental table 1)^6, 8, 10, 16, 17, 23^ yet lower sensitivity of the orofacial region to thermal stimuli compared to healthy participants in other studies^14^. However, QST studies outside the orofacial region in BMS patients, such as leg and arm extremities, report mixed results, including higher^8^, lower^21^, and non-differing^15^ HPTs compared to healthy participants.

These conflicting extra-trigeminal QST findings could be due to the cyclical nature of spontaneous pain in BMS^4^. Therefore, we focus on BMS type I because patients experience little to no pain in the morning and as the day progresses their pain increases peaking in the afternoon^1, 11^. Thus, if spontaneous pain is related to changes in sensitization, we would expect different QST results at different times of the day in BMS type I. However, this temporal evaluation of somatosensory responses in BMS type I remains unknown.

In the current study, we examined psychophysical responses to thermal and pressure stimuli on the face and forearm in the morning and afternoon in BMS type I patients and healthy participants to address how time of day affects somatosensory responses in BMS type I. We also collected pain diaries from BMS patients across eight days to illustrate the cyclical nature of pain in this BMS sample. We hypothesized that compared to healthy participants, BMS patients have higher pain sensitivity, specific to the orofacial regions, during the afternoon.

## Materials and Methods

### Overview of data collection

All research procedures were granted approval by the Institutional Review Board of the University of Maryland, Baltimore. Three groups were used for analysis.

BMS participants were asked to complete a two-day experimental session comprised of 3 days of pain diaries followed by test day 1, and test day 2 culminating with 3 more days of pain diaries. We required visit 2 to be within 9 days after visit 1. An option of a one-day experimental session was offered in order to reduce scheduling conflicts and increase enrollment. The one-day experimental session comprised of 3 days of pain diaries followed by test day 1, and culminating with 4 more days of pain diaries.

### Participants

#### Recruitment criteria

BMS patients were recruited following diagnosis at the Oral Medicine Program at the University of Maryland School of Dentistry (led by TFM), where complete dental and oral health examinations were performed. A working diagnosis of BMS was based on a chief complaint of pain or burning in the oral mucosa and/or tongue and exclusion of other known causes of oral burning-like pain. If BMS patients were taking topical medications or in the transition of weaning off a systemic medication, to start a new one, we asked them to come in when they had completely weaned off of the medications and would test them prior to their transition into a new medication regimen.

Additionally, all healthy participants were recruited through campus-wide flyer advertisement and were free of any chronic pain conditions, psychiatric illness, local oral or systemic disease, and salivary dysfunction.

##### Exclusion criteria for all studie

subjects unable or refusing to sign consent for any part of the testing; any chronic pain conditions; claustrophobia; pacemaker; weight over 300 lbs; daily regimen of opiates; excessive alcohol use as measured on the AUDIT^20^ on hormone replacement therapy within the last 30 days. For BMS cohorts, if participants were on a systemic medication regimen they were excluded.

#### Enrollment

Informed consent was obtained from each participant according to the Declaration of Helsinki. Altogether, we enrolled 51 total post-or peri-menopausal female participants: 18 BMS patients and 33 healthy participants (table 1).

**Table 1:** number of participants and ages. ^s.d.^standard deviation.

| All postmenopausal women | Current study |  | Previous study | Concurrent study | Group totals |  |
| --- | --- | --- | --- | --- | --- | --- |
|  | healthy group 1 | BMS group 1 | healthy group 2 | healthy group 3 | Total healthy | Total BMS |
| Number of participants | 11 | 18 | 10 | 12 | 33 | 18 |
| Age (s.d) | 60.8<br>(±8.8) | 60.6<br>(±5.7) | 54.9<br>(±7.6) | 52<br>(±3.78) | 55.7<br>(±7.66) | 61.3<br>(±6.4) |

The healthy control group consisted of data from three separate studies: healthy control group 1 (n=11) was enrolled in the current protocol with the BMS patients; healthy control group 2 (n=10) was obtained from a previous study from our laboratory with identical methods for some of the QST procedures; and healthy participant group 3 (n=12) was concurrently run study in our laboratory with identical methods for some of the QST procedures.

BMS and control group 1 volunteers presented themselves for a two-day or a one-day experimental session. For the two-day experimental session, we randomized whether a participant would experience a morning (AM) or an afternoon (PM) QST session on the first day and on the second day participants were assigned the opposite time of day (AM/PM) for QST testing. For example, if a participant was given afternoon QST on the first day, they would have QST in the morning of the second day. For participants who could not commit to two testing days we offered a one-day experimental session, where we randomized whether a participant would experience a morning or an afternoon QST session.

### Diaries

BMS patients were given an 8-day paper diary to track their oral burning pain intensity and unpleasantness. For the two day visit, they completed the diaries for 3 consecutive days prior to the laboratory visit, during the two day visit, and for 3 consecutive days after the visit. For the one day visit, they completed the diaries for 3 consecutive days prior to the laboratory visit, during the one day visit, and for 4 consecutive days after the visit. Participants were asked to rate their burning pain intensity on a scale of 0-10, with 0 meaning ‘none’ and 10 meaning ‘as bad as you can image’ and unpleasantness on a scale of 0-10, with 0 meaning ‘not bothersome’ and 10 meaning ‘extremely bothersome’, at 5 different time-points: wakeup, 10am, 2pm, 6pm and bedtime each day.

### Thermal testing

Thermal heat and cold stimuli were delivered to the forearm and face via a 27mm in diameter Medoc Pathway CHEPS Peltier thermode with a heating rate of 70°C/sec and a cooling rate of 40°C/sec (Pain & Sensory Evaluation System, Medoc Advanced Medical Systems Ltd., Ramat Yishai, Israel). Four thermal tests where administered: three tests on the forearm (temperature threshold, “levels”, and “ratings” (not reported here), testing) and one test on the face (temperature threshold testing). The thermode was repositioned along the forearm and cheek after each stimulus to avoid temporal summation.

#### Forearm temperature threshold testing

Participants received a warmth detection threshold (WDT) test, where the temperature increased from a baseline temperature of 32°C at a rate of 1°C/sec until mouse click. Then, they received a cool detection threshold (CDT) test, where the temperature decreased from 32°C at a rate of 1°C/sec until mouse click. In both WDT and CDT, participants were asked to click the mouse when they first detected a change in temperature. We then tested heat pain threshold (HPT), where temperature increased from 32°C at a rate of 1.5°C/sec until mouse click. Participants were asked to press the mouse as soon as the temperature first became painful. WDT, CDT, and HPT were each performed three times and were calculated as the average temperature across the three trials.

#### Face Temperature threshold testing

Following the forearm temperature threshold testing procedures, participants received WDT, CDT, and HPT tests with temperature stimuli presented three times for each test as explained above; however this time the thermode was placed on the left cheek. The temperature increased (WDT, HPT) or decreased (CDT) from a baseline temperature of 32°C at a rate of 1°C/second until mouse click when they first detected a change in temperature (WDT, CDT) and as soon as the temperature first became painful (HPT). WDT, CDT, and HPT were each performed three times and were calculated as the average temperature across the three trials.

#### Forearm levels testing

Subsequently, a “levels” test was administered to the forearm where participants received a series of heat stimuli delivered in ascending order of target temperatures: 35°, 35°, 39°, 41°, 43°, 45°, 47°, and 49°C. We varied the ramp rates to each target temperature in order to maintain the same ramp time of 1.6 seconds with each heat stimulus, from the baseline temperature of 32°C. Target temperatures were sustained for 6 seconds, so the total heat stimulus duration including ramps was 9.2 seconds. A 20 second inter-stimulus interval between target temperatures allowed the participant to input their rating for the presented target temperature. Participants rated pain intensity on a numerical rating scale (NRS) of 0 (no pain) to 10 (extremely intense pain) and pain unpleasantness on an NRS of 0 (not bothersome) to 10 (extremely bothersome pain).

Subsequently, we averaged the first pain intensity and unpleasantness ratings of the first two temperature exposures (35°C and 35°C) to obtain a single value for 35°C and leaving us with 7 total temperature exposures in the morning and afternoon (14 temperatures total). Next, we created a pain intensity <u>Total comparison</u>comprised of the AM and PM pain intensity ratings averaged together to obtain a single value per temperature (7 values total) per group (BMS vs healthy) (see statistical analyses section for more details). The same procedure was followed to create a pain unpleasantness <u>Total comparison per group</u>(BMS vs healthy).We also created an <u>AM comparison</u>, the 7 temperature stimulations in the AM are averaged together to get a single value per temperature (7 values total) per group (BMS vs healthy). The same procedure was followed to create a <u>PM comparison per group</u>(BMS vs healthy). Additionally, the same procedure was followed to create an <u>AM comparison</u> and <u>PM comparison</u> pain unpleasantness rating per group (BMS vs healthy).

### Pressure pain threshold testing

Bilateral pressure pain thresholds (PPT) were obtained using a Wagner Force Dial tm FDK 20 /FDN Series Push Pull Force Gage pressure algometer (20 lb x .25 lb; 10 kg x 100 gr). Participants received pressure stimuli at four locations of the body: left thumbs, elbows, temporalis, and masseter muscles (repeated three times in that sequential order) and then right thumbs, elbows, temporalis, and masseter muscles (repeated three times in the listed sequential order). Participants were asked to raise a hand when the pressure first became painful and the pressure at that instant was recorded in kilograms.

As explained above, participants were presented with pressure a total of 6 times (three times on the left and three times on the right side of the face (temporalis and masseter) and extremities (thumbnail and elbow)). Subsequently, we created a PPT <u>Total comparison</u>comprised of the AM and PM 12 pressure exposures (left and right temporalis) averaged together for each group (BMS vs healthy). We created a PPT <u>AM comparison</u> the six AM pressure exposures (left and right temporalis) averaged for each group (BMS vs healthy). The same procedure was followed for the PM comparison. Additionally, we created a PPT <u>AM vs PM comparison</u> using the six PPT AM averages and six PPT PM averages for each group (BMS vs healthy).

### Statistical Analysis

For each type of QST, the following four comparisons were performed (supplemental table 2 and table 2): 1. <u>Total comparison</u>: Mann-Whitney U test between groups (healthy vs BMS) of the average of both time points (AM and PM). For these analyses, all participants in the BMS and healthy groups were included, whether they had data from AM alone, PM alone, or both AM and PM, in which case an average was taken. 2. <u>AM comparison</u>: Mann-Whitney U test between group analyses of data taken from each subject at the AM time point. 3. <u>PM comparison</u>: Mann-Whitney U test between group analyses of data taken from each subject at the PM time point. 4. <u>AM vs PM comparison</u>: Wilcoxon signed-rank test within group analyses of AM and PM time points within BMS and healthy groups. We also performed an area under the curve (AUC) with respect to ground analysis of pain intensity and unpleasantness ratings for the levels test. AUC was calculated according to the literature following the same grouping as listed above^17^. Because of the small sample size in each group non-parametric tests were performed.

**Table 2:** comparisons and the statistical tests used

| Comparison type | Comparison name | Description | Statistical test |
| --- | --- | --- | --- |
| Between group | Total | [AM + PM of BMS] vs [AM + PM of Healthy] | Mann-Whitney U test |
| Between group | AM | AM of BMS vs AM of Healthy | Mann-Whitney U test |
| Between group | PM | PM of BMS vs PM of Healthy | Mann-Whitney U test |
| Within group | AM vs PM | AM BMS vs PM BMS<br>Or AM Healthy vs PM Healthy | Wilcoxon signed-rank test |

Additionally, separate models were run due to the variation in sample size in each group.

#### Diaries

Mean, median, and range of BMS pain intensity ratings were reported for each of the 8 days. A Friedman test was used to compare average pain intensity for 5 time points in a day across 8 days, followed by a Dunn’s post-hoc test for multiple comparisons. No diaries were collected from healthy participants as ratings of burning mouth pain and unpleasantness were presumably zero and therefore no comparisons were made for healthy participants.

## Results

### Demographics

All participants were peri-or post-menopausal women. BMS participants had an age range of 47 to 74 years (mean 61, SD ± 6). Seventy-seven percent of BMS participants were Caucasian, six percent African American, six percent Asian, and eleven percent mixed race. Healthy participants had an age range of 43 to 73 years (mean 56, SD ± 8). Seventy-nine percent of healthy participants were Caucasian, seventeen percent African American, and four percent Asian.

### Diaries

Pain intensity ratings of BMS patients were significantly higher as the day progressed from wake to bedtime (p<0.0001) (Figure 1). Post-hoc analyses revealed pain intensity ratings increased from baseline by 3.3 at 2pm (p=0.0268), 3.6 at 6pm (p<0.001) and bedtime (p=0.0003) compared to wake time. In addition, there were no significant differences in pain intensity ratings across each individual time point and the 8 days of diary recordings; for example, there was no significant difference in pain rating of wake time across the 8 days, no significant difference at 9am across 8 days, and so on.

**Figure 1:**
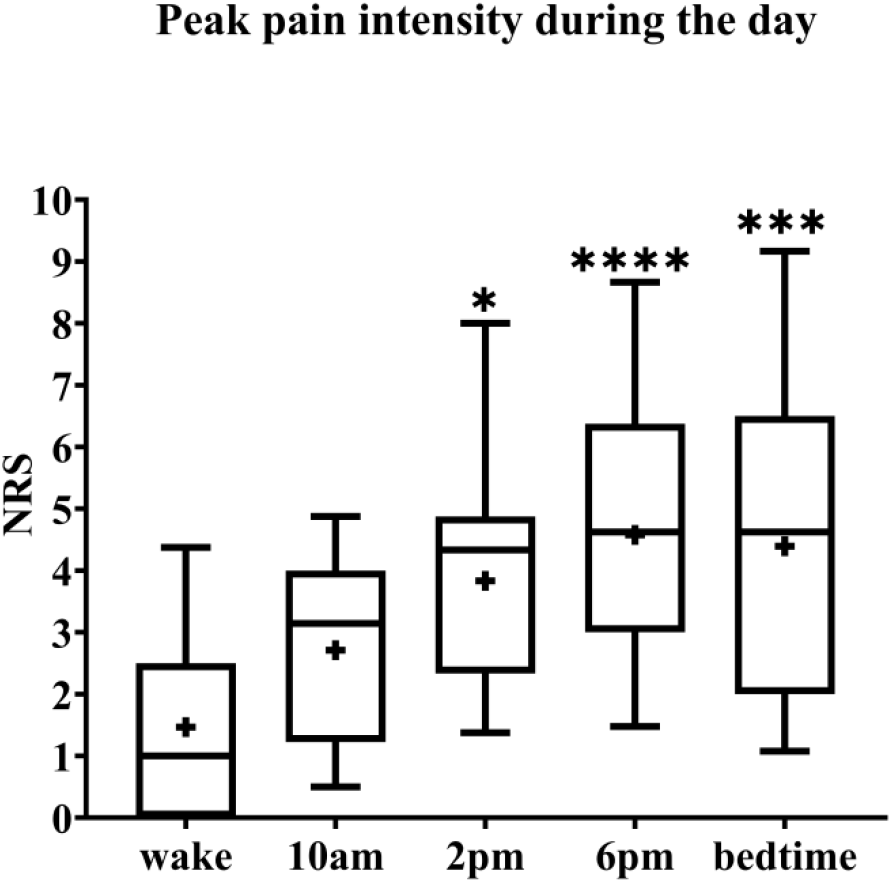
Pain intensity ratings of BMS patients across 8 days. The box spans the interquartile range, whiskers represent the full range, horizontal line within each box mark the median, and the + represents the mean. *p=0.02, ***p=0.0003, and ****p<0.0001 compared to wake. NRS: numerical rating scale.

### Thermal testing in BMS versus healthy participants

### Warmth detection thresholds

#### Face

In the <u>Total comparison,</u> BMS patients had significantly lower WDTs than healthy participants by 2.2°C (p=0.0494) (figure 2). There were no significant differences in the <u>AM comparison</u>, <u>PM comparison</u>, or <u>AM vs PM comparison</u> within or between groups (figure 3).

**Figure 2:**
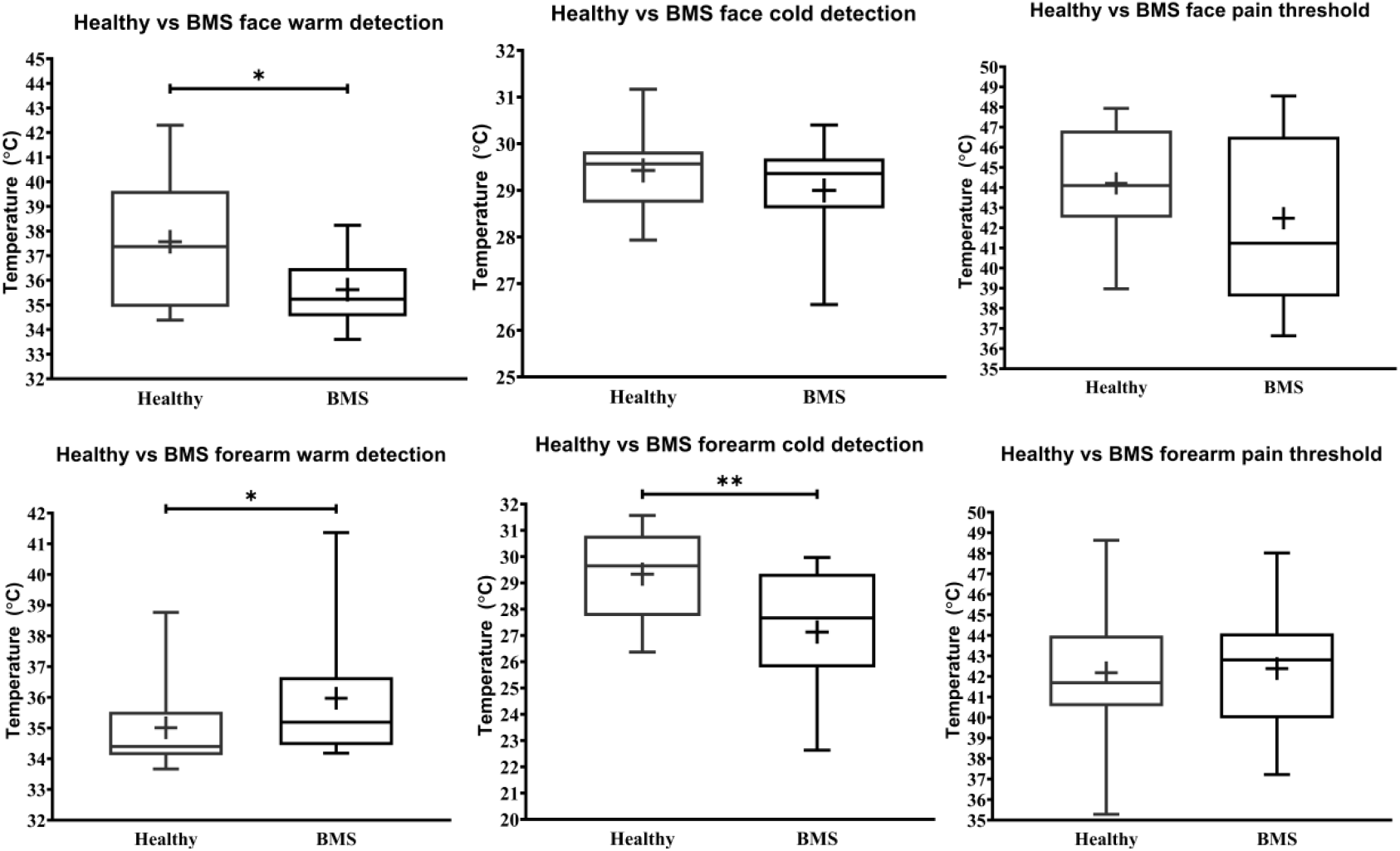
<u>Total comparison</u> of temperature detection and pain threshold in BMS patients compared to healthy participants. WDT, CDT, HPT are shown consecutively in order of exposure to the face (top) and forearm (bottom). *p<0.05 and **p<0.005.

**Figure 3:**
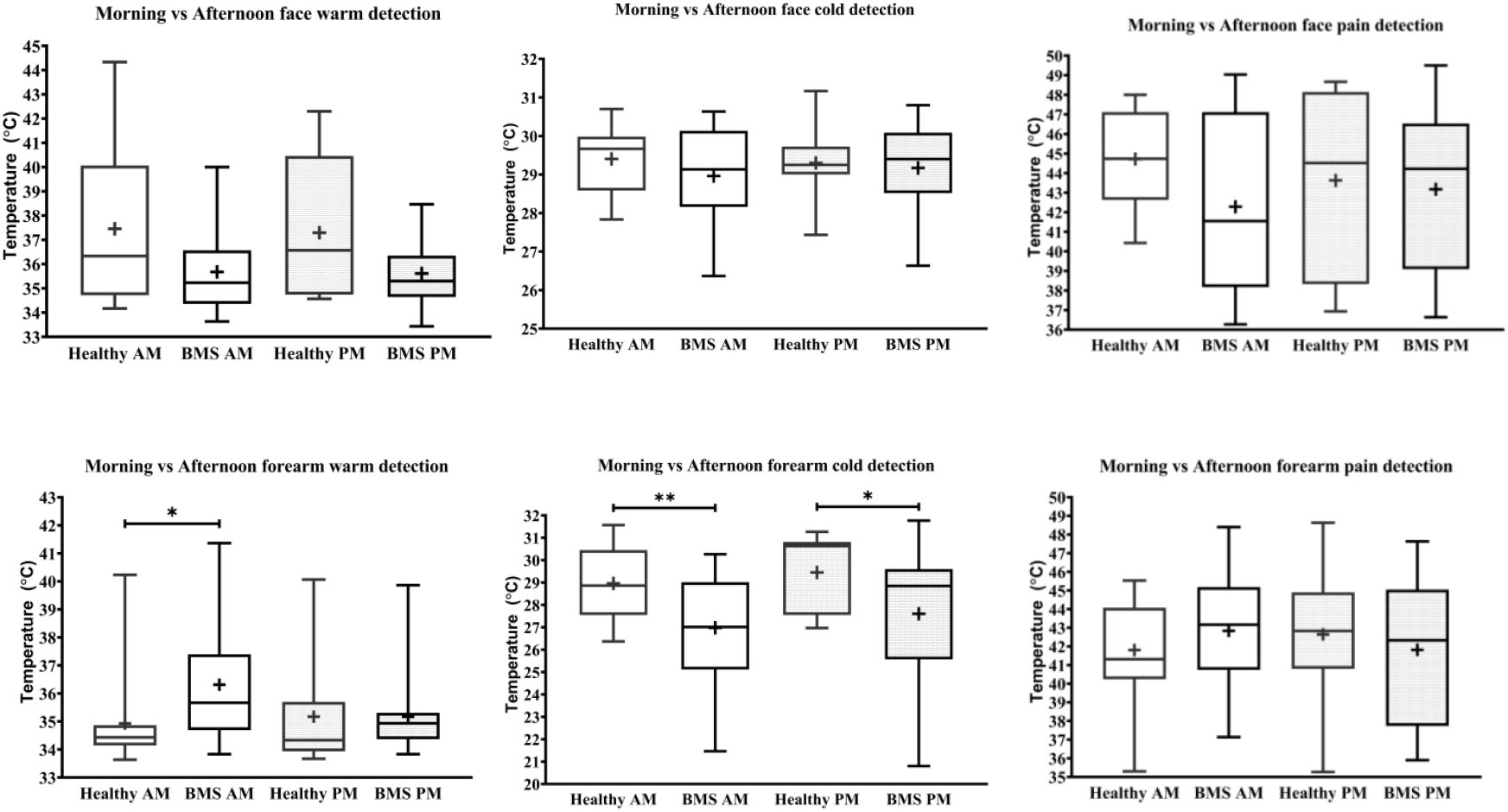
Morning versus afternoon temperature detection and pain threshold in BMS patients and healthy participants. Face (top) and forearm (bottom) measures. ^*^p<0.05, ^**^p<0.005.

#### Forearm

In the <u>Total comparison,</u> BMS patients had significantly higher WDTs compared to healthy participants by 0.8°C (p=0.0172) (figure 2). In the <u>AM comparison</u>, BMS patients had significantly higher WDTs compared to healthy participants by 1.3°C (p=0.0113). There were no differences in the <u>PM comparison</u> or in the <u>AM vs PM comparison</u>.

### Cold detection thresholds

#### Face

BMS patients had no significant differences in CDT compared to healthy participants in the <u>Total comparison</u> (figure 2). There were no significant differences in <u>AM comparison</u>, <u>PM comparison</u>, or <u>AM vs PM comparison</u> within or between groups (figure 3).

#### Forearm

In the <u>Total comparison,</u> BMS patients had significantly lower CDTs compared to healthy participants by 2°C (p=0.0021) (figure 2). In the <u>AM comparison</u>, BMS patients had significantly lower CDTs compared to healthy participants by 1.9°C (p=0.0096). In the PM comparison, BMS patients had significantly lower CDTs compared to healthy participants by 1.7°C (p=0.0397). In the <u>AM vs PM comparison, t</u>here were no differences within groups (figure 3).

### Heat pain thresholds

#### Face

BMS patients had no significant difference in HPT compared to healthy participants in the <u>Total comparison</u> (figure 2). There were no significant differences in <u>AM comparison</u>, <u>PM comparison</u>, or <u>AM vs PM comparison</u> within or between groups (figure 3).

#### Forearm

BMS patients had no significant difference in HPT compared to healthy participants in the <u>Total comparison</u> (figure 2). In the <u>AM comparison</u>, <u>PM comparison</u>, or <u>AM vs PM comparison</u>, there were no significant differences within or between groups (figure 3).

### “Levels” forearm pain testing BMS versus healthy participants

In the <u>Total comparison,</u> BMS participants had significantly higher pain intensity at 35°C by 2.583 pain ratings (p=0.0006), 39°C by 1.583 pain ratings (p=0.0386), 41°C by 1.417 pain ratings (p=0.0412), 43°C by 3 pain ratings (p=0.0167), 45°C by 2 pain ratings (p=0.0146), 47°C by 2.75 pain ratings (p=0.0201) (figure 4). There were no differences in the AUC for pain intensity or unpleasantness <u>Total comparison</u>.

**Figure 4:**
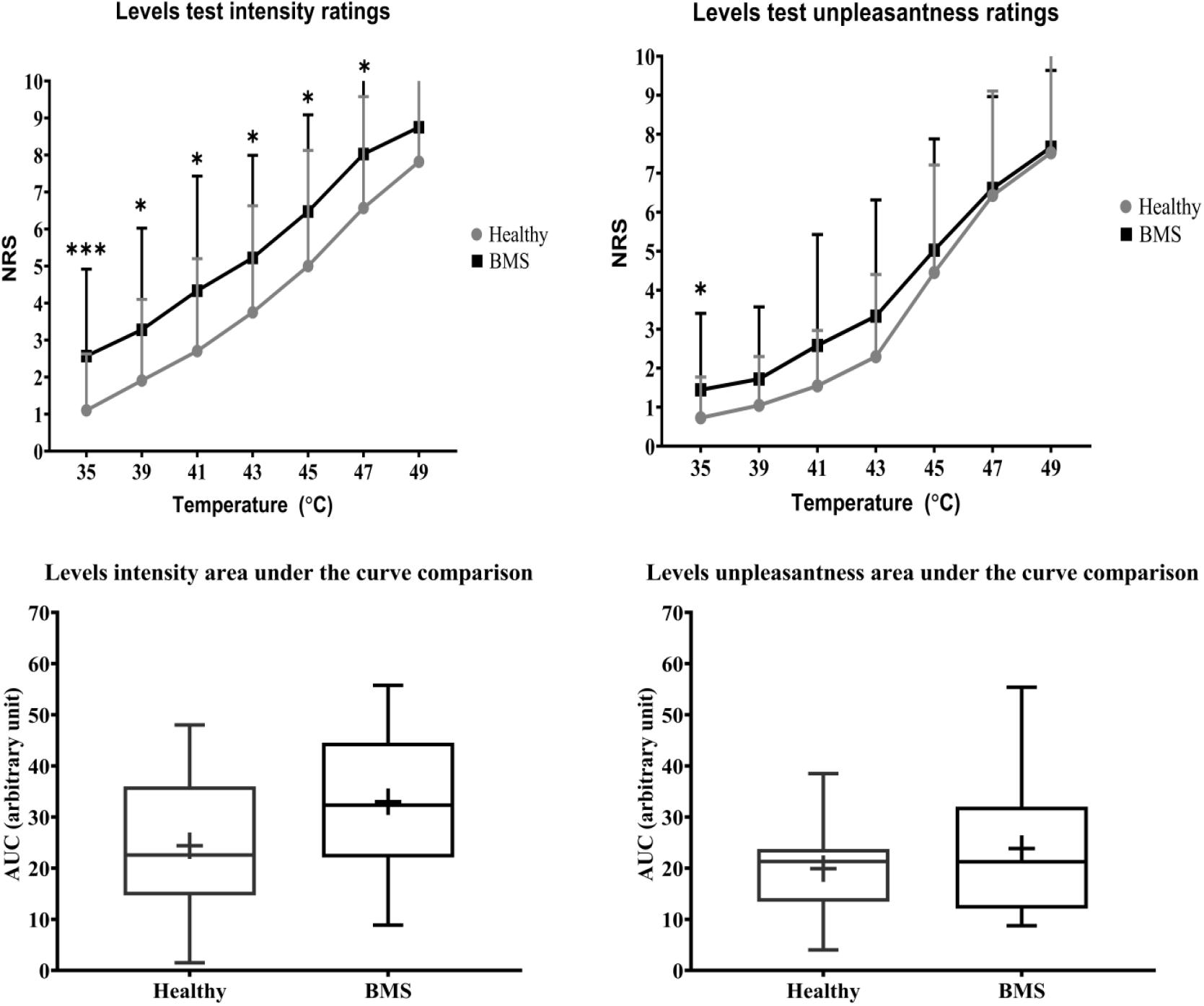
<u>Total comparison</u> of “levels” forearm responses in BMS patients relative to healthy participants. Averaged intensity (left) and unpleasantness (right) responses per temperature and the respective standard deviation. The boxplot (bottom) shows the overall effect of healthy and BMS patients on pain intensity and unpleasantness as AUC. *p<0.05. ***p<0.001.

In the <u>PM comparison</u>, BMS participants had significantly higher pain intensity ratings at 35°C by 1.75 pain ratings (p=0.0206). There were no significant differences in the <u>AM comparison</u> nor in the <u>AM vs PM comparison</u>. There were no differences in AUC for <u>AM comparison</u>, <u>PM comparison</u>, or <u>Am vs PM comparison</u>.

In addition, BMS had significantly higher pain unpleasantness ratings at 35°C by 0.583 pain ratings (p=0.0112) in the <u>Total comparison</u>. In the unpleasantness AUC <u>AM vs PM comparison</u>, there were no significant differences in between groups (figure 5).

**Figure 5:**
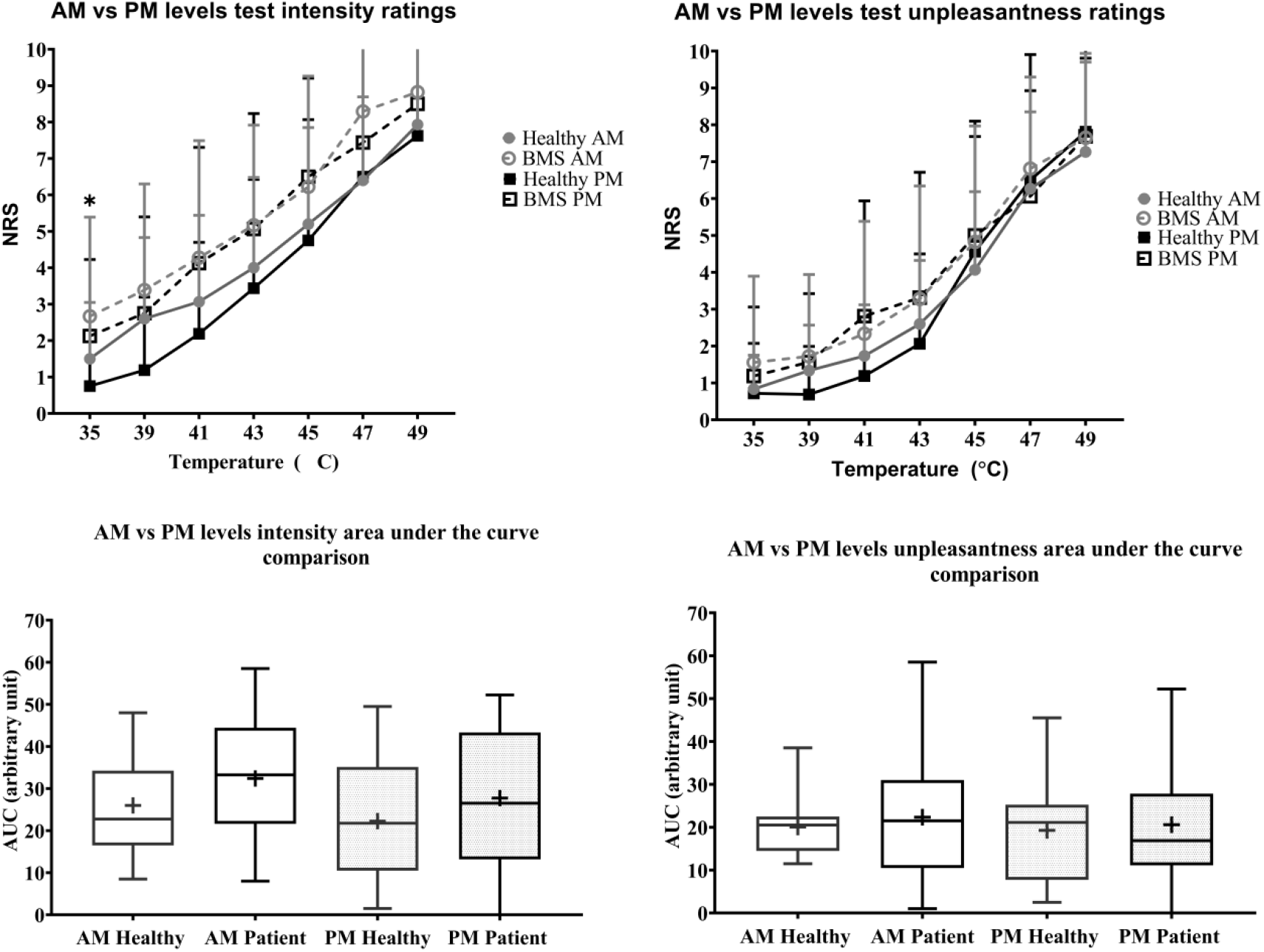
Morning versus afternoon “levels” forearm responses in BMS patients and healthy participants. Averaged intensity (left) and unpleasantness (right) responses per temperature in the morning (AM) and afternoon (PM) and the standard deviation. The boxplot (bottom) shows the overall effect of healthy and BMS patients on pain intensity and unpleasantness as AUC.*p<0.05 for BMS vs Healthy in the PM.

### Pressure testing in BMS versus healthy participants

In the <u>Total comparison,</u> BMS patients had no significant differences in PPTsof the masseter, temporalis, thumbnail, and elbow compared to healthy participants (figure 6). In the <u>AM vs PM comparison</u>, BMS patients had significantly lower within group PPTs of the masseter in the afternoon by 0.34kg (W=79, p=0.0413) (figure 7). There were no significant differences in PPTs of the masseter, temporalis, thumbnail, and elbow in the <u>AM comparison</u>, <u>PM comparison</u>between groups.

**Figure 6:**
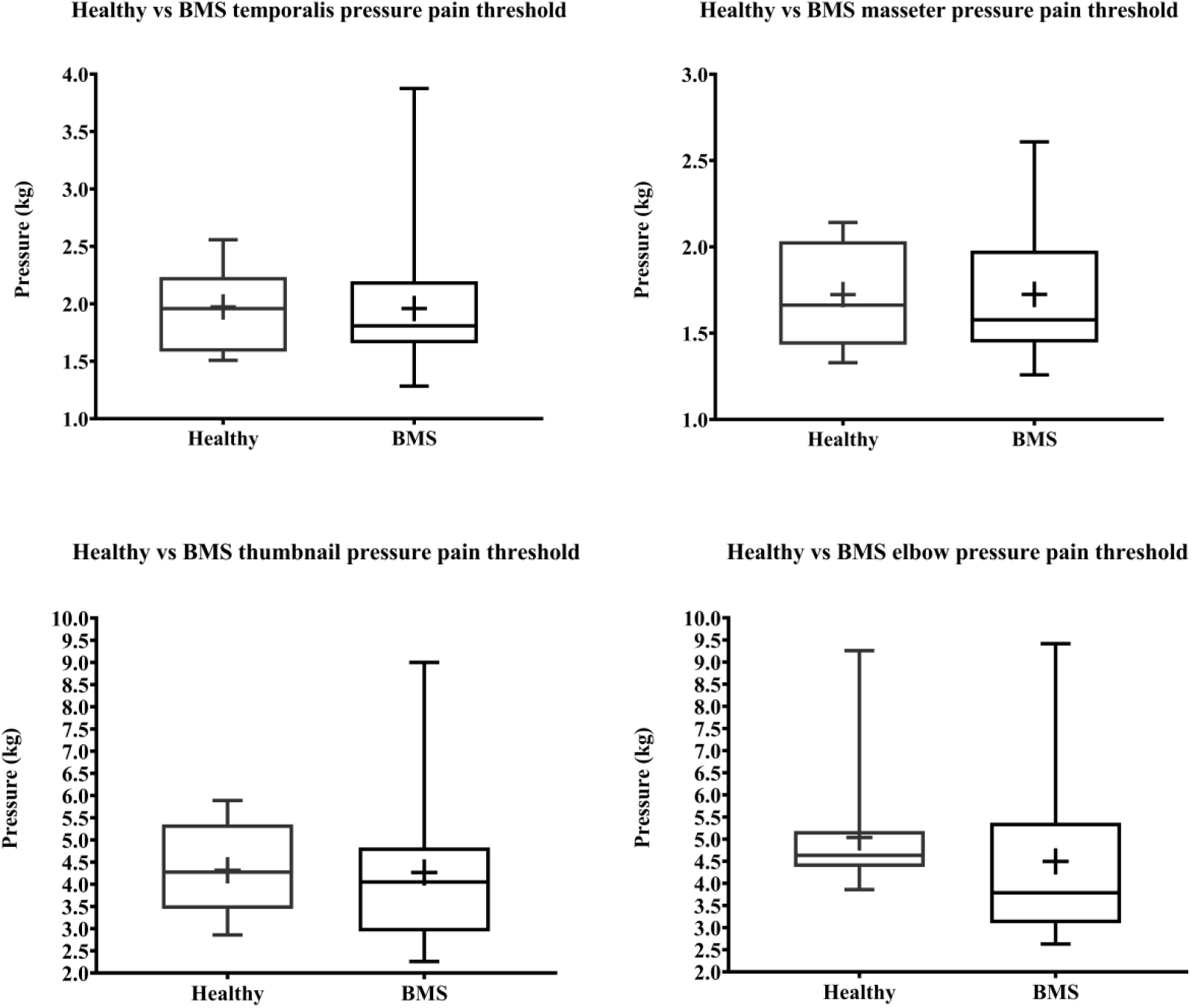
<u>Total comparison</u> of pressure pain thresholds for face (top) and extremity (bottom) of BMS patients and healthy participants.

**Figure 7:**
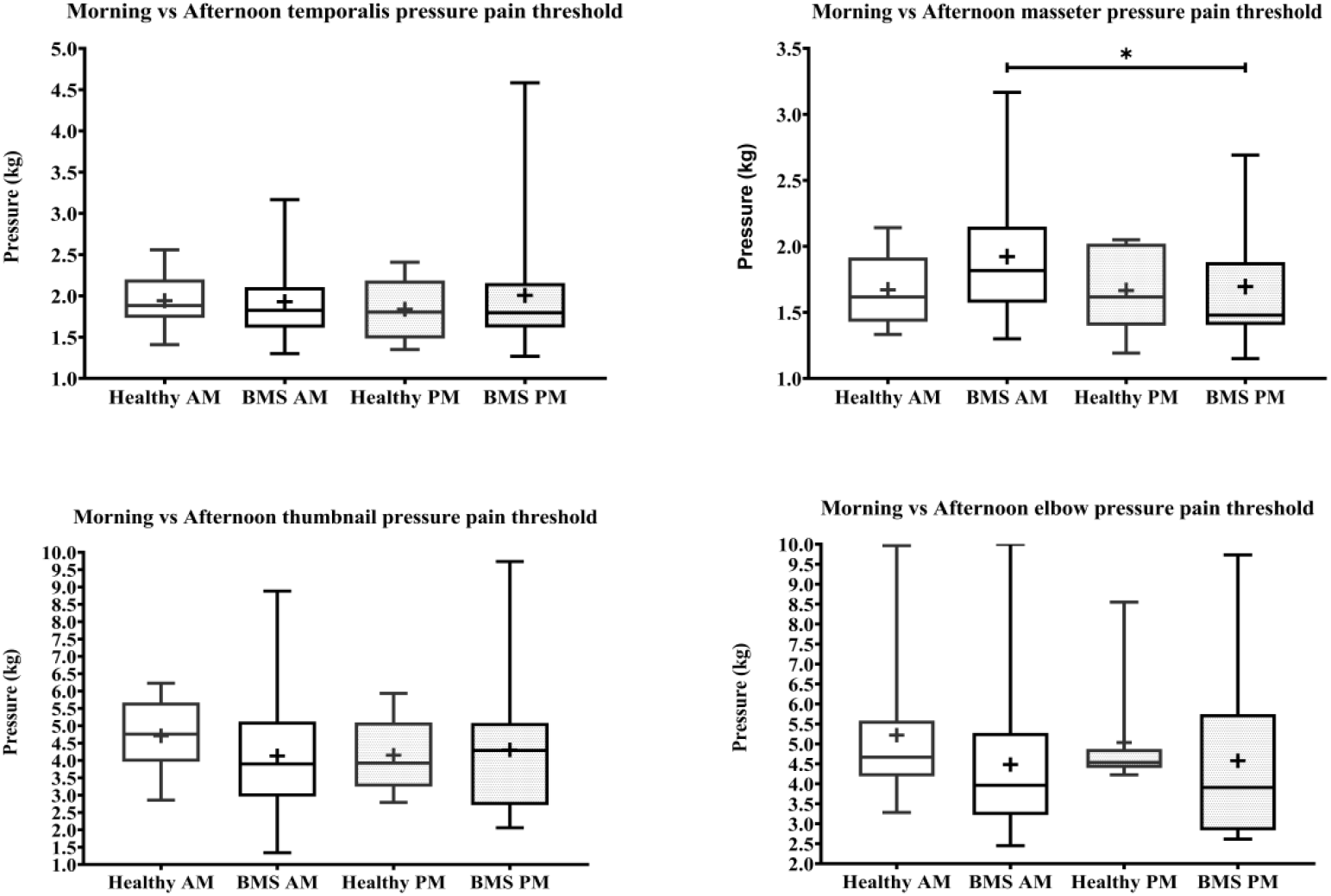
Morning versus afternoon pressure pain thresholds comparisons for face (top) and extremity (bottom) of BMS patients and healthy participants.*p<0.05.

## Discussion

In this study, we investigated whether sensory sensitivity of the orofacial region and the forearm was affected by time of day and thus the presence of ongoing pain in BMS type I patients compared to healthy participants. Our overall hypothesis that compared to healthy participants, BMS type I patients have higher pain sensitivity, specific to the orofacial regions and in the afternoon, was not supported. Our main findings showed that time of day has a significant effect on the spontaneous orofacial pain as quantified by pain diaries. However, there were no differences in experimental thermal pain to the orofacial region, and only experimental pressure pain to the masseter showed as significant effect. Instead, we found a significant time of day effect to the experimental thermal exposure to the forearm, with BMS patients displaying less sensitivity to both cold and warm temperature detection, but higher pain sensitivity to heat at low temperatures. This is the first study to compare morning to afternoon QST pain measures in BMS patients compared to healthy subjects.

BMS type I is characterized as a burning sensation that is not present upon waking, but which develops in the late morning and progresses during the waking hours, with the greatest intensity of discomfort in the evening^1^. Based on diary records, we found that spontaneous pain intensity became significantly higher as the day progressed, and the ratings were mostly consistent for each participant across the 8 days of testing. This confirmed the pattern of ongoing pain in BMS type I, i.e., higher pain ratings in the afternoon compared to morning and that pain sensation is present every day^1, 11^. We also expected that as the day progresses BMS patients would have increased within group orofacial pain and pain sensitivity to other stimuli such as thermal and pressure. This within group expectation was only supported by thea single finding that BMS participants are more sensitive to pressure applied to the masseter muscle in the afternoon when compared to morning. There were no effects of time of day within the BMS group for any other QST measure on any region tested.

We did not find time of day differences between BMS patients and healthy participants in orofacial thresholds assessed by morning and afternoon comparisons of WDT, CDT, HPT, nor temporalis muscle PPT. Even though the lack of differences could simply be due to our low sample size, Mo et al. found no differences in PPTs of the hand, tongue, chin, or lip between groups with a comparable sample size of 25 BMS and 19 healthy participants^14^. Thus, enhanced orofacial pain sensitivity in BMS patients may be independent on time of day.

We also found no time of day differences in the extremities assessed by thumbnail and elbow PPT comparisons between BMS and healthy participants. Similar to our findings Watanabe et al, did not find differences in PPTs of the forearm, although they did report increased forearm mechanical pain sensitivity ^21^. Therefore, a possible interpretation is that BMS does not affect pain evoked by pressure but instead affects mechanical sensitivity of the extremities.

Central sensitization has been suggested as a potential mechanism for the presence of pain in other body regions of BMS patients^4, 9, 12^. We found that BMS patients have lower sensitivity to non-noxious thermal stimulation displayed by the higher WDTs and lower CDTs in the morning at the forearm relative to healthy participants, which does not support a role of central nervous system changes leading to widespread hypersentivity. Instead, the hyposensitivity to cold and warm temperature on the forearm may be due to hypervigilance to their ongoing spontaneous BMS pain as opposed to experimentally evoked thermal stimulation, a phenomenon previously observed in other chronic pain conditions^7, 15^. Hypervigilance is as an enhanced state of sensory sensitivity accompanied by an exaggerated search for threatening information, which may in turn exacerbate the pain experience^15, 22^. Thus, as their BMS pain spontaneously starts to surface in the morning, patients may develop a pain-specific “hypervigilance” to their orofacial pain as a result of continual effort to detect BMS related painful sensations of the orofacial region even in the presence of non-painful cold and warm stimulation on the body. In essence, it can be interpreted that their hypervigilance to the onset of BMS related pain distracts them from the experimentally evoked thermal perception which reflects in lower sensitivity to external innocuous stimuli in the morning. This finding is further supported by the lower CDTs in the afternoon on the forearm in BMS compared to healthy participants and the lower PPTs of the masseter within the BMS patient group in the afternoon when compared to morning.

We found some unexpected outcomes in BMS patients. HPTs tested on the face and forearm in BMS patients did not differ from those in healthy participants. Prior literature on HPTs show conflicting results, including higher, lower, and non-differing HPTs compared to healthy participants in the orofacial region as well as the extremities^6, 8, 10, 16, 21, 23^. It was also unexpected that there were no overall differences in CDTs on the face between groups, regardless of time of the day. Prior literature on CDTs and WDTs are also inconsistent including no differences in WDTs on body regions^8, 21, 23^, lower CDTs on the tongue^8^, and slightly higher intraoral CDTs in BMS compared to healthy participants^22^. Further research is necessary to fully address the contradictory findings in the BMS field and investigate potential mechanisms underlying individual differences between BMS type I patients. We interpret that some of the inconsistent findings in the field could be due to the lack of consideration of the cyclicity of the BMS type I and suggest that incorporating morning and afternoon comparisons can help reduce the variability.

The findings in the present study should be interpreted in light of some limitations. First, sample size was relatively small. However, in order to obtain a homogeneous sample, we excluded BMS patients with comorbidities, on opiates, and under hormone replacement therapy and we only included patients who met strict criteria based on clinical diagnosis of BMS type I, which drastically decreased the number of eligible patients. Second, we were limited by the types of tests we could perform in BMS patients. No intra-oral sensory testing was performed and we only performed the levels test on the arm in order to prevent triggering BMS discomfort to patients by applying suprathreshold stimuli to the face. Third, we did not have a direct measure to infer central sensitization in BMS patients.

In conclusion, warm and cold processing is impaired in BMS type I patients, which could suggest hypervigilance toward clinically relevant pain that results in reduced sensitivity to innocuous stimuli applied to distal body areas. Despite clear increase in spontaneous pain, we saw limited time-of-day dependent effects on QST measures. Subsequent studies should consider potential mechanisms underlying individual differences in BMS type I patients, and investigate the impact of pain and other sensory sensitivities in brain signaling in order to further understand BMS symptomatology.

## Data Availability

N/A

## Acknowledgements

The authors have no financial conflicts of interest to declare. We thank Massieh Moayedi, Man-Kyo Chung, Marjorie Toensing, and Andrea R. Gray for feedback on the manuscript. We would also like to thank Sharon Varlotta, Dianna Weikel, Shariq Khan for help with participant scheduling, Mariya Prokhorenko, Janusiya A. Muthulingam, Marc Rangel, and Samuel Krimmel for help with equipment and data collection, Luma Samawi, Taylor Duckworth, Drew Resh, Daniela Loebl, and Brandon Boring for helping with data management. Additionally, we would like to thank all our funding sources for making this work possible: NIDCR, NIH F31 DE027622-01A1, NIH 2T32NS063391-11A1, and NIGMS, NIH R25-GM55036 to JPS; NIDCR, NIH R21 DE023964-01A1 and 3R21DE023964-02S1 to DAS.

## Supplemental tables

**Supplemental Table 1:** Literature review findings. ^↑^BMS higher than healthy BMS. ^↓^lower than healthy. ^−^no difference between groups.

|  | Orofacial or body | Temp or pressure? | warm | cold | heat | mechanical pain sensitivity | Mechanical detection threshold |
| --- | --- | --- | --- | --- | --- | --- | --- |
| Grushka et al, 1987 | orofacial | temp | N/A | N/A | ↓ | N/A | N/A |
| Kaplan et al, 2011 | orofacial | temp | N/A | N/A | - | N/A | N/A |
| Mo et al, 2015 | orofacial | both | ↑ | ↓ | ↑ | - | N/A |
| Watanabe et al, 2019 | both | both | N/A | N/A | ↓ body | ↑ body (<6months BMS) | ↑ tongue (<6months BMS) |
| Yang et al, 2019 | both | both | ↑ face | ↑ face | ↓ face - body | ↓ face | ↑ face |
| Honda et al, 2019 | body | temp | N/A | N/A | ↑ body | N/A | N/A |

**Supplemental Table 2:**
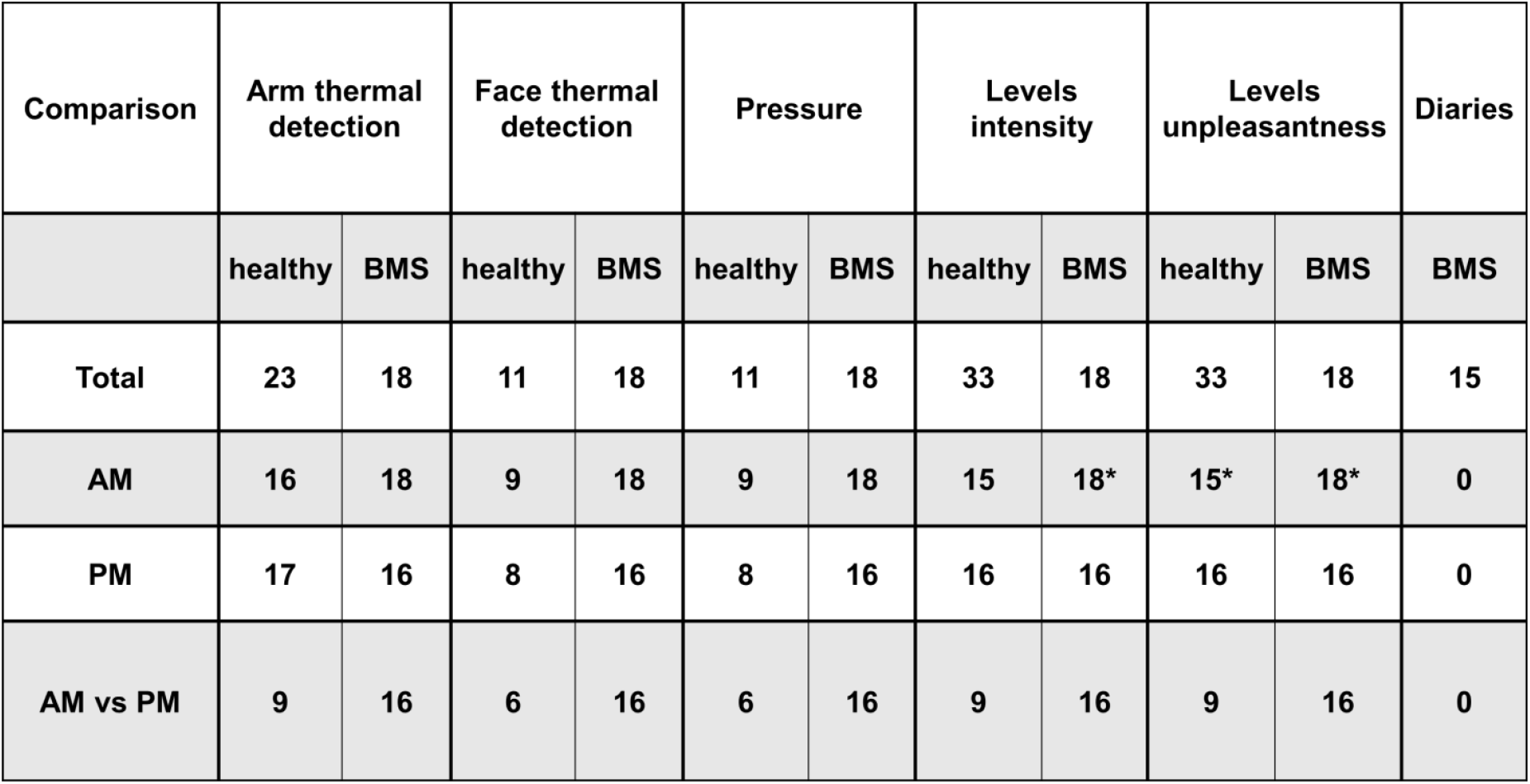
number of participants in each test and comparison. *represents exceptions where number of participants was lower by one participant than stated in table due to missing data: at * AM levels intensity had an n=17 BMS at 47°C and at 49°C; at * AM Levels unpleasantness had an n=14 healthy and an n=16 BMS at 47°C; and an n=17 BMS at 45°C and 49°C.

